# Socioeconomic Status, Biological Aging, and Memory in a Diverse National Sample of Older US Men and Women

**DOI:** 10.1101/2021.06.04.21258370

**Authors:** Justina F. Avila, Indira C. Turney, Precious Esie, Jet M. J. Vonk, Vanessa R. Weir, Daniel W. Belsky, Jennifer J. Manly

## Abstract

**Importance:** Exposure to socioeconomic disadvantage is associated with rapid cognitive aging. Biological aging, the progressive loss of system integrity that occurs as we age is proposed as a modifiable process mediating this health inequality.

**Objective:** To test the hypothesis that socioeconomic disparities in cognitive aging in older adults is explained by accelerated biological aging.

**Design:** Observational cohort study.

**Setting:** U.S. Health and Retirement Study DNA-methylation sub-study.

**Participants:** N=3,648 (49% male) adults aged 50-100 (M=70, SD=10) with DNA methylation data.

**Exposures:** Socioeconomic status (SES) was measured from years of education and household wealth. The extent and pace of biological aging were quantified using three DNA-methylation measures: PhenoAge, GrimAge, and DunedinPoAm.

**Main Outcomes and Measures:** Cognitive aging was measured from repeated longitudinal assessments of immediate and delayed word recall. Latent growth curve modeling estimated participants’ level of memory performance and rate of decline over 2-11 follow-up assessments spanning 2-20 years. Models were estimated to assess whether the relationship between SES and memory trajectories was mediated by biological aging.

**Results:** Older adults with lower SES had lower memory performance, faster decline and exhibited accelerated biological aging (SES β=.33, 99% CI[.30-.36], biological-aging measure effect size associations ranged from .08 to .20). Accelerated biological aging was associated with decreased memory performance and faster memory decline (effect-size range .03 to .18). Higher SES was associated with slower biological aging for White and Black men and women, but not Latinx participants. The relationship between biological aging measures and memory was weaker for Black and Latinx participants compared with White people. In mediation analysis, biological aging accounted for 3-9% of the SES-memory gradient in White participants. There was no evidence of mediation in Black or Latinx participants.

**Conclusions and Relevance:** Among a national sample of older adults, DNA-methylation measures of biological aging were associated with memory trajectories in White, but not Black or Latinx older adults. These results challenge the assumption that DNA-methylation biomarkers of aging that were developed in primarily White people can quantify aging processes affecting cognition in Black and Latinx older adults.

---

Aging and socioeconomic status (SES) are two important determinants of late-life cognitive health. Physicians often regard age and SES as immutable factors rather than quantities that could be modified to improve patient health. However, both may represent promising targets for intervention. Moreover, they may be connected.

The biological process of aging involves the progressive loss of system integrity, causing decreased resilience of cellular networks and organ systems, ultimately leading to disease and death.^1^ The process of biological aging begins in early life^2^ and manifests in variable rates of decline in organ-system integrity from as early as young adulthood.^3^ In midlife adults, faster-paced decline in organ-system integrity is associated with signs of brain aging and cognitive decline.^4–6^ Aging-related changes in brain integrity (e.g., cortical thinning, hippocampal atrophy) are accompanied by decline in cognitive performance.^7^

Low SES, commonly measured from wealth, income, occupational status, and/or educational experience, is associated with an earlier onset and faster pace of aging-related changes in the brain.^8–12^ The mechanism by which SES inequalities affect health may be through acceleration of the aging process via repeated adaptation to stressors that cause cumulative wear and tear on the body’s system and increases vulnerability to multiple disease processes.^13–18^ Until recently, we have not had a way of comprehensively measuring these processes. DNA-methylation biomarkers that quantify molecular alterations that occur with aging provide promising tools for measuring SES inequalities in aging. Several proposed measures indicate more advanced biological aging in individuals with greater socioeconomic disadvantage.^19–23^ Some of these measures have also been associated with cognitive performance in small samples.^20,23,24^ However, it is unclear whether DNA-methylation patterns mediate the relationship between SES and late-life cognitive health.

Most research on DNA-methylation biomarkers of aging has focused on White, Educated, Industrialized, Democratic (WEIRD) samples. Further, population subgroup differences in the pathways that link SES to cognition may be present at the intersection of race/ethnicity and sex/gender groups, which is critical to consider for potential intervention on SES or biological aging. The overall goal of the current study was to determine whether DNA-methylation biomarkers are associated with memory trajectories in a representative national sample of older adults and to examine whether these measures of biological aging mediate the relationship between SES and late-life memory trajectories similarly across racial/ethnic-sex/gender subgroups.

## Methods

### Participants

The Health and Retirement Study (HRS) is a nationally representative longitudinal study of Americans over age 50, designed to examine the health, social, and economic factors associated with aging.^25^ Data collection began in 1992, with the initial cohort born between 1931-1941 (aged 51-61 in 1992). In 1993, a second cohort was added consisting of individuals born before 1924 (aged ≥70 in 1993). Two additional cohorts were added in 1998 to address age gaps and create a representative sample of those aged >50. These additional cohorts consist of individuals born between 1924-1930 and 1942-1947, respectively. Since then, a new cohort of individuals aged 51-56 has been continually added to the HRS sample every 6 years (2004, 2010, 2016). The HRS oversamples Black and Hispanic/Latino/a/x/e (Latinx, hereafter) participants to improve reliability of estimates.^26^ Additional details of the HRS sample may be found elsewhere.^27^

Participants are followed-up every two years to complete core interviews. These core interviews consist of content including (but not limited to): demographics, assets and income, physical conditions and treatment, health behaviors, cognitive function, among others.^25^ In 2016, HRS participants were asked to consent to a venous blood draw, of which samples from 9,934 participants were collected (65% completion rate among eligible cases).^28^ DNAm assays were performed on a sub-sample of these participants (n=4,104); about 98% passed quality control checks (n=4,018). This DNA-methylation sample is representative of the entire HRS sample. Participants were excluded from the current analyses if they were missing data on race/ethnicity (n=2) and years of education (n=19). The remaining sample (n=3,997) included 1,161 non-Latinx White men, 1,501 non-Latinx White women, 227 non-Latinx Black men, 432 non-Latinx Black women, 225 Latinx men, 331 Latinx women, and 47 men and 73 women who identified their ethnicity as non-Latinx and race as other.

### Measures

#### Measures of Biological Aging

There is no gold standard measure of biological aging; several methods have been proposed based on different biological measurements and analytic strategies.^29^ We focused on measurements available in the HRS database for which there was consistent evidence of association with SES and cognitive aging: two DNA-methylation clocks, PhenoAge^30^ and GrimAge,^31^ and the DunedinPoAm^20^ pace of aging measure. The three aging measures are described in detail in the **Supplemental Methods**.

In short, the PhenoAge and GrimAge clocks estimate the age at which a person’s mortality risk would be approximately normal in their respective reference samples: PhenoAge was developed in the US NHANES III and InCHIANTI studies,^30^ GrimAge was developed in the Framingham Heart Study Offspring cohort.^31^ Both clocks were developed using 2-step approaches that involved modeling physiological parameters (clinical chemistries and complete blood count data for PhenoAge, blood proteins for GrimAge) and mortality risk. Clock-ages that are older than the chronological age of the person being measured indicate an advanced state of biological aging; younger clock-ages indicate delayed aging.^32^ For analysis, HRS participants’ DNA-methylation-clock ages were regressed on their chronological ages and residual values were computed. These residual values, referred to as “age acceleration residuals” were standardized to mean=0, SD=1.

DunedinPoAm estimates a person’s pace of aging, the rate of decline in system integrity that occurs with advancing chronological age. DunedinPoAm was developed by analyzing longitudinal change in 18 biomarkers tracking multi-organ-system integrity in a birth cohort followed from ages 26-38.^20^ Values are interpretable as years of physiological change occurring per 12-month calendar interval in healthy adults. DunedinPoAm values >1 indicate a faster than normal pace of aging; values <1 indicate a slower pace of aging. For analysis, values of DunedinPoAm were standardized to mean=0, SD=1.

#### Race/Ethnicity and Sex/Gender

HRS collected information about race/ethnicity by (i) asking participants whether they were Hispanic or Latino; and (ii) asking participants to classify themselves racially as White, Black, Asian, American Indian, Alaska Native, or Pacific Islander. Sex/gender was determined as male or female (it is unknown whether individuals reported their biological sex or their preferred gender identification).^33^

#### Socioeconomic Status

An SES composite was created based on years of education and wealth. Years of education was measured by the highest self-reported completed grade of school. We calculated birth cohort-based education standardized scores (z-scores) by subtracting each participant’s years of education by the mean education for their birth cohort and dividing by its SD. Wealth was measured by *total wealth* available in the HRS RAND income and wealth imputation dataset.^34^ *Total wealth* at each study visit was adjusted for inflation and transformed using the inverse hyperbolic sine.^35^ Z-scores were created for wealth based on 5-year age groups (i.e., 50-54, 55-59, 60-64, etc.) at each study wave to adjust for age and period effects. Z-scores were then averaged across study waves for each participant to represent average wealth during the study. Lastly, education and wealth z-scores were averaged to represent the SES composite.

#### Memory Performance

Memory was assessed via the immediate and delayed recall scores of the CERAD^36^ 10-item word list memory test. Only data from Wave 3 (1996) or later were used because this was the first wave the 10-item memory test was administered. We restricted longitudinal data to visits that respondents were aged >50. Raw scores on immediate and delayed recall trials were converted into z-scores using the means and SDs at baseline. Composite scores were computed by averaging these z-scores at each occasion.^37^

### Statistical Analyses

Memory trajectories from 1996 to 2016 were modeled as a latent growth curve with time centered at the 2016 visit, indicating the amount of time, in years from 2016, that each respondent participated in previous sessions. Thus, intercepts indicate memory performance when DNA samples were collected, and slopes indicate the average rate of decline throughout the study. Age at 2016 visit (B-spline) and initial study wave were included covariates. Intercept and slope estimates were saved for each participant and used as outcomes in subsequent models to avoid multicollinearity issues.^38^ We estimated a series of models to evaluate independent relationships between each DNA-methylation indicator, SES, and memory intercept and slope. Next, formal mediation analyses were conducted, separately for each DNA-methylation indicator. For these models, we specified paths between (1) SES and the DNA-methylation indicator, (2) the DNA-methylation indicator and memory outcomes, and (3) SES to the memory outcomes. The indirect effect was calculated as the product of the path between SES and DNA-methylation indicator and the path between DNA-methylation indicator and memory.^39^ Standardized parameter estimates are reported.

Each model was first estimated across the entire sample. To examine racial/ethnic-sex/gender subgroup differences, we employed known-class mixture models with racial/ethnic-sex/gender subgroups as known classes. This known grouping variable is incorporated as a moderator variable, allowing model parameters to vary as a function of membership in the identified groups. Multiple-group models were only estimated for White, Black, and Latinx men and women (N=3,877), since sample sizes of the other racial/ethnic-sex/gender subgroups were too small for this type of analysis.

All analyses were performed using Mplus version 8.5.^40^ Models were estimated using a Bayesian mediation approach with diffuse prior distributions, which can handle smaller sample sizes and produces credible intervals with probabilistic interpretation rather than confidence intervals.^41,42^ Sensitivity analyses were conducted that included sampling weights for the 2016 Venous Blood Study to obtain population-based estimates. Both p-values and confidence intervals were used to determine statistical significance.^43^ To decrease the likelihood of type I error due to multiple comparisons we used a p-value of .001 (i.e., 99% confidence interval).

## Results

Characteristics of the full sample and the six racial/ethnic-sex/gender subgroups are presented in Table 1. Participants had an average of 4.2 (SD=3.1) visits with cognitive data. Black men demonstrated the lowest 2016 memory performance and White women had the highest. Compared with White men and women, Black women had steeper memory decline.

**Table 1.** Sample Characteristics

| Characteristics | Entire Sample<br>(N=3,648) | White Men<br>(N=1,160) | White Women<br>(N=1,498) | Black Men<br>(N=227) | Black Women<br>(N=432) | Latinx Men<br>(N=225) | Latinx Women<br>(N=331) |
| --- | --- | --- | --- | --- | --- | --- | --- |
| Age, mean (SD) | 69.89 (9.6) | 71.45 (9.5) | 70.98 (9.8) | 67.07 (8.4) | 66.31 (8.7) | 66.13 (8.0) | 65.75 (9.0) |
| Years of Education, mean (SD) | 13.30 (2.6) | 13.71 (2.5) | 13.44 (2.3) | 12.50 (2.7) | 12.72 (2.6) | 9.88 (4.6) | 9.11 (4.7) |
| Wealth, mean (SD)<sup>a</sup> | 425.21 (1152.4) | 528.88 (1094.5) | 513.86 (1459.1) | 116.12 (254.6) | 92.39 (240.3) | 136.62 (242.0) | 160.27 (423.9) |
| SES, mean (SD) | -0.02 (0.8) | 0.26 (0.6) | 0.18 (0.7) | -0.33 (0.6) | -0.32 (0.6) | -0.69 (0.8) | -0.75 (0.9) |
| Epigenetic Clocks |  |  |  |  |  |  |  |
| PhenoAge Residual, mean (SD) | -0.01 (6.91) | 0.61 (6.7) | -0.66 (6.9) | 0.24 (7.3) | 0.19 (7.3) | 0.75 (6.4) | -0.13 (6.1) |
| GrimAge Residual, mean (SD) | 0.03 (4.8) | 1.63 (4.6) | -1.73 (4.3) | 3.17 (5.0) | 0.35 (4.8) | 1.57 (4.2) | -1.54 (3.8) |
| DunedinPoAm | 1.08 (0.1) | 1.07 (0.1) | 1.06 (0.1) | 1.11 (0.1) | 1.09 (0.1) | 1.09 (0.1) | 1.06 (0.1) |
| Memory Performance, mean (SD) | -0.46 (0.02) | -0.49 (0.04) | -0.27 (0.03) | -0.96 (0.1) | -0.77 (0.1) | -0.67 (0.1) | -0.85 (0.1) |
| Memory Decline, mean (SD) | -0.04 (0.01) | -0.04 (0.01) | -0.04 (0.01) | -0.04 (0.01) | -0.06 (0.01) | -0.03 (0.01) | -0.05 (0.01) |

Abbreviations: SD, standard deviation; SES, socioeconomic status composite variable (z-score)
<sup>a</sup> Values reported are the Total Wealth variable divided by 1000.

SES was strongly associated with 2016 memory performance (β=0.44 [0.39, 0.46]) but weakly associated with memory decline (β=0.09 [0.03, 0.12]). Higher SES was associated with higher memory level and slower decline across racial/ethnic-sex/gender subgroups; however, this relationship was stronger for Black and Latinx compared with White women and men (Figure 1).

**Figure 1.**
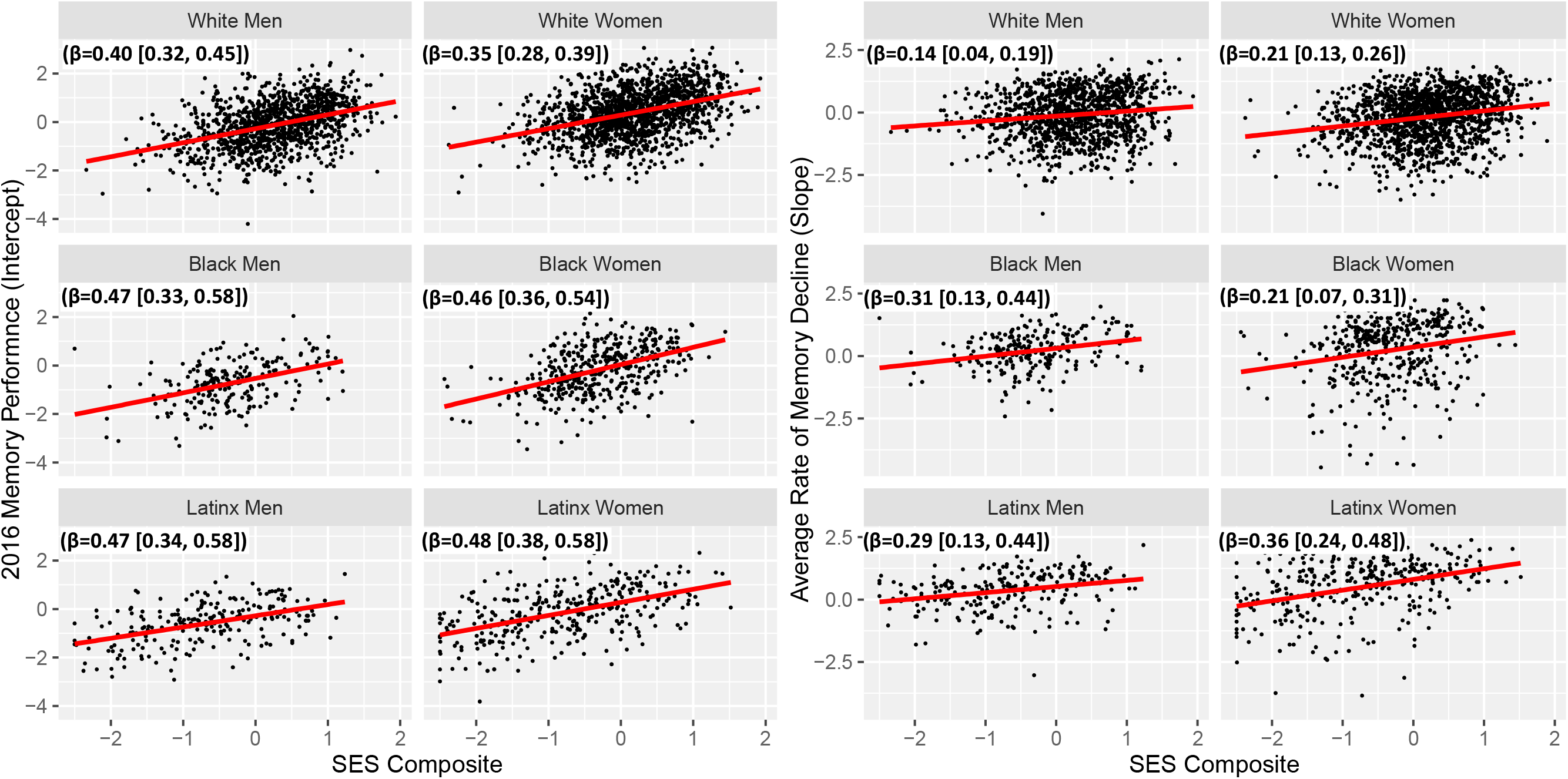
Socioeconomic Gradients in Memory Level and Decline Across Racial/Ethnic-Sex/Gender Subgroups.

Participants’ DNA-methylation clock ages were highly correlated with their chronological ages (PhenoAge r=0.73; GrimAge r=0.83). DunedinPoAm was not correlated with chronological age (r=0.02). Across race/ethnicity-sex/gender groups, PhenoAge indicated that Latinx men had the most-advanced aging and Latinx and White women had the most delayed aging. GrimAge and DunedinPoAm indicated that Black men had the fastest biological aging and White and Latinx women had the slowest aging.

An SES gradient in biological aging was noted in the entire sample, where higher SES was associated with slower aging for each DNA-methylation measure (PhenoAge β=-0.08 [-0.12, - 0.04]; GrimAge β=-0.20 [-0.24, -0.16]; DunedinPoAm β=-0.17 [-0.21, -0.13]. The SES-biological aging gradient was similar across White and Black men and women for GrimAge and DunedinPoAm; for PhenoAge the gradient was present only for White women (Figure 2). The SES gradient was not present with any of the DNA methylation measures for Latinx men and women.

**Figure 2.**
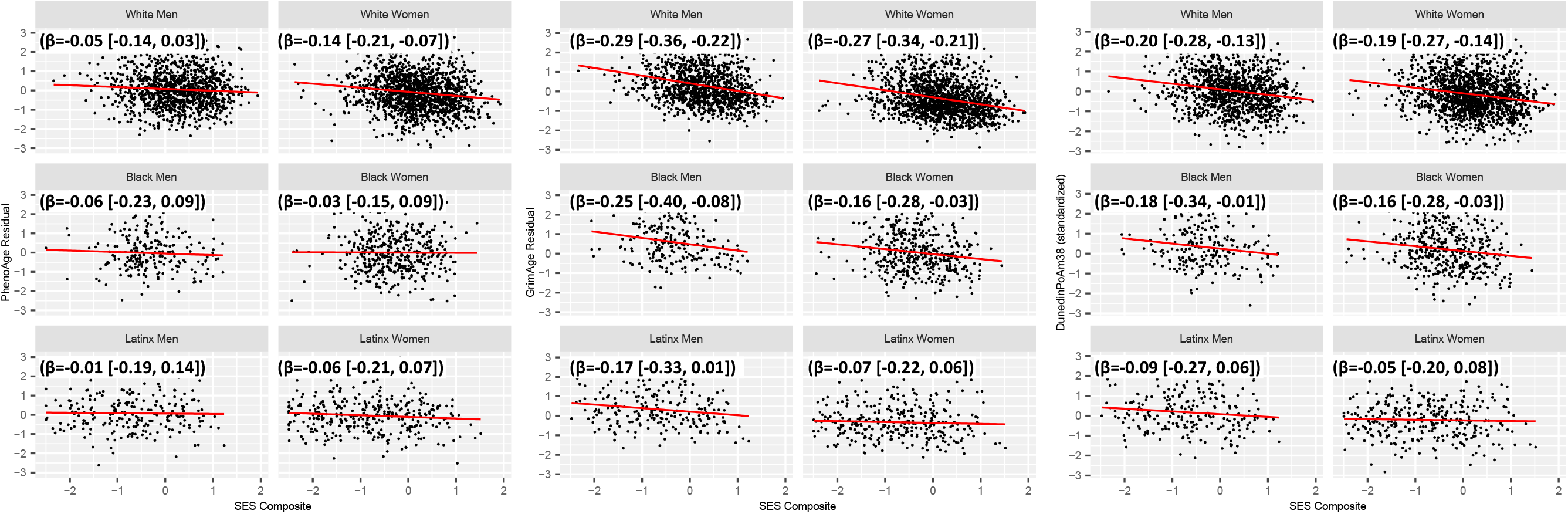
Socioeconomic Gradients in DNA methylation Measures of Aging Across Racial/Ethnic-Sex/Gender Subgroups.

Overall, individuals with accelerated biological aging demonstrated lower 2016 memory performance (PhenoAge β=-0.08 [-0.13, -0.04]; GrimAge β=-0.25 [-0.29, -0.21]; DunedinPoAm β=-0.16 [-0.19, -0.11]). More-advanced PhenoAge (β=-0.04 [-0.09, -0.001] and GrimAge (β=-0.06 [-0.11, -0.02]) and faster DunedinPoAm (β=-0.08 [-0.11, -0.02]), were associated with faster memory decline. Associations of DNA-methylation measures of aging with memory performance varied across race/ethnic and sex/gender groups (Figure 3). PhenoAge was only associated with 2016 memory performance for White women. Older GrimAge and Higher DunedinPoAm values were associated with lower 2016 memory performance and faster memory decline for White men and women, but not Black or Latinx men or women.

**Figure 3.**
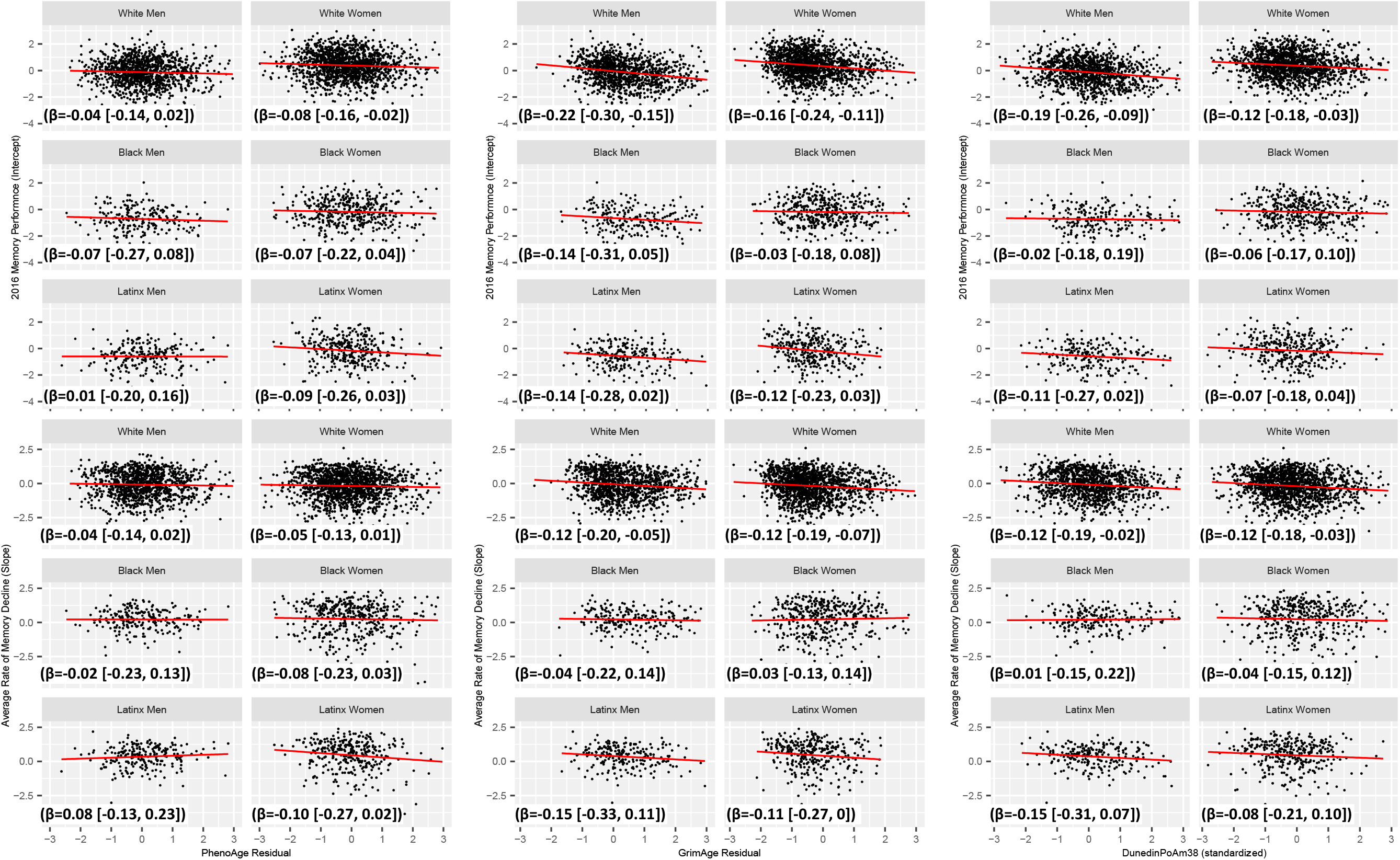
Relationship Between DNA methylation Measures of Aging and Memory Level and Decline Across Racial/Ethnic-Sex/Gender Subgroups.

Mediation models were estimated for each DNA-methylation measure separately. Indirect effect estimates are presented in Table 2. Each DNA-methylation measure partially mediated the SES gradient on 2016 memory performance. Proportion mediated (PM) estimates indicated that GrimAge (PM=8%) conveyed a larger portion of the SES gradient than DunedinPoAm (PM=4%) or PhenoAge (PM=0.3%). GrimAge (PM=11%) and DunedinPoAm (PM=13%) partially mediated the SES-memory decline gradient. No mediation effects were noted for the multiple-group PhenoAge mediation model. GrimAge partially mediated the SES gradient in 2016 memory for White men (PM=8%) and women (PM=9%) and the SES-memory decline gradient for White men (PM=19%) only, but not for other groups. Similarly, DunedinPoAm partially mediated the SES-2016 memory gradient only for White men (PM=6%) and women (PM=2%), as well as the SES-memory decline gradient only for White men (PM=14%) and women (PM=7%). Across mediation models, none of the DNA-methylation measures were reliably associated with memory performance among Black and Latinx men and women. There were no reductions in the relationship between SES and memory level and decline when DNA-methylation measures were included in the models. Results from sensitivity analyses that included HRS sampling weights did not substantively differ from the unweighted results.

**Table 2.** Indirect Effect Estimates for Mediation Models (Standardized Estimates [99% Confidence Intervals])

| Indirect Effect | SES → PhenoAge →<br>2016 Memory | SES → GrimAge →<br>2016 Memory | SES → DunedinPoAm →<br>2016 Memory |
| --- | --- | --- | --- |
| Entire Sample | 0.004 (0.001, 0.007) | 0.03 (0.02, 0.04) | 0.02 (0.01, 0.02) |
| White Men | 0.001 (-0.002, 0.01) | 0.03 (0.01, 0.05) | 0.02 (0.01, 0.04) |
| White Women | 0.004 (-0.10, 0.01) | 0.02 (0.001, 0.03) | 0.01 (0.001, 0.02) |
| Black Men | 0.002 (-0.01, 0.03) | 0.01 (-0.03, 0.06) | -0.01 (-0.05, 0.02) |
| Black Women | 0.001 (-0.01, 0.02) | -0.01 (-0.03, 0.01) | -0.002 (-0.03, 0.01) |
| Latinx Men | 0 (-0.02, 0.02) | 0.01 (-0.02, 0.06) | 0.01 (-0.01, 0.05) |
| Latinx Women | 0.003 (-0.01, 0.03) | 0.01 (-0.01, 0.03) | 0.001 (-0.01, 0.03) |

| Indirect Effect | SES → PhenoAge →<br>Memory Decline | SES → GrimAge →<br>Memory Decline | SES → DunedinPoAm →<br>Memory Decline |
| --- | --- | --- | --- |
| Entire Sample | 0.002 (-0.001, 0.006) | 0.01 (0.001, 0.02) | 0.01 (0.004, 0.02) |
| White Men | 0.001 (-0.001, 0.01) | 0.02 (0.003, 0.05) | 0.02 (0.004, 0.03) |
| White Women | 0.002 (-0.007, 0.01) | 0.02 (0, 0.04) | 0.02 (0.003, 0.03) |
| Black Men | 0 (-0.02, 0.02) | -0.01 (-0.06, 0.04) | -0.01 (-0.05, 0.02) |
| Black Women | 0.002 (-0.01, 0.02) | -0.01 (-0.04, 0.01) | 0.001 (-0.02, 0.02) |
| Latinx Men | 0 (-0.02, 0.04) | 0.02 (-0.02, 0.07) | 0.01 (-0.01, 0.06) |
| Latinx Women | 0.003 (-0.01, 0.03) | 0.003 (-0.01, 0.03) | 0.001 (-0.01, 0.02) |

Abbreviations: SES, socioeconomic status composite variable (z-score)

## Discussion

DNA-methylation biomarkers of aging are emerging as a powerful approach for understanding how social exposures, such as SES, get under the skin to shape health in aging. We tested how three DNA-methylation measures representing the extent and pace of biological aging related to the socioeconomic gradient in memory trajectories in a nationally representative sample of older adults. There were two main findings. First, socioeconomic gradients in biological aging were present in Black and White men and women; those with more wealth and more years of school exhibited less-advanced biological aging. Second, biological aging mediated a portion of the socioeconomic gradient in memory level and decline for White men and women, but not for Black or Latinx men and women.

Our finding that higher SES was associated with less-advanced/slower biological aging in White and Black people in the US extends observations from studies of European samples.^20–23^ This result builds confidence that the PhenoAge and GrimAge clocks and DunedinPoAm pace of aging, which were developed in primarily White samples, provide valid measures of aging in Black people. In Latinx people, however, these DNA-methylation measures were not sensitive to biological impact of SES variation.

Results from our analysis of memory functioning raise questions, especially in the use of DNA-methylation clocks relate to cognitive aging in Black and Latinx individuals. DNA-methylation clocks and related measures, such as the DunedinPoAm pace of aging, represent a cutting-edge approach to quantifying biological aging in epidemiologic and clinical studies. So far, few studies have described how these measurements relate to the process of cognitive aging. In the large US-representative sample we analyzed, older adults with more-advanced biological aging demonstrated lower memory performance and faster memory decline than those of the same chronological age with less-advanced/slower biological aging. This finding expands on observations from smaller, primarily White European and New Zealand samples^20,23,24^ to establish a link between the extent and pace of biological aging and later-life memory decline. However, in tests of association within race/ethnic-sex/gender groups not represented in those prior studies, specifically Black and Latinx men and women, the DNA methylation measures were not consistently associated with memory trajectories. Little attention has been paid to potential group differences in the relationship between DNA-methylation biomarkers and aging outcomes. While it is possible that biological aging may play more of a role in cognitive aging for Whites compared with non-Whites, these findings highlight the need for rigorous consideration of these novel measures of biological aging in minoritized populations. Racially-patterned lifecourse social experiences may shape epigenetic processes differently across racial/ethnic groups and such profiles may not be adequately represented with these current DNA-methylation measures.

Even among White men and women, the DNA-methylation measures mediated only a small fraction of the SES gradient in aging-related memory decline. The relationship between SES and late-life cognitive functioning is more complex than the simple mediation model examined in this study.^44–46^ It is possible that the primary impact of biological aging is on other intermediary factors (e.g., cardiovascular disease) on the pathway linking SES to memory trajectories. Other molecular pathways, not measured by the DNA-methylation measures available in the HRS study may be more salient biological links between SES and late-life memory.

A limitation of the current study is the relatively small number of non-White HRS participants with DNA-methylation measures. Larger samples would have greater power to detect small effects of DNA-methylation measures in mediation models, if present.^47^ Another limitation is that each DNA-methylation measure was obtained at a single point in time. While individuals who showed a steeper decline in memory throughout the study demonstrated more advanced biological aging in 2016, analyses cannot rule out reverse causation (i.e., poor memory performance contributing to advanced biological aging).

## Conclusions

Among a nationally representative sample of older adults in the US, individuals with more-advanced/faster biological aging as measured by the GrimAge DNA-methylation clock and DunedinPoAm pace of aging had lower scores on memory tests and faster decline in memory functioning over time. GrimAge and DunedinPoAm also indicated more advanced and faster aging in lower SES older adults. However, biological aging measures mediated only small fractions of the SES gradient in aging-related memory decline. Moreover, associations of biological aging measures with memory trajectories were consistent only in Whites but not Black or Latinx people. DNA-methylation biomarkers of aging have potential to provide surrogate endpoints for interventions that aim to prevent aging-related functional decline, including cognitive decline, but lose that potential if their utility is limited to White people.^48^ Minoritized communities face disproportionate burden of cognitive decline and dementia. Our results suggest that current DNA-methylation biomarkers of aging do not quantify aging processes affecting cognition in Black and Latinx older adults as well as they do in White older adults. If so, a next generation of measures is needed.

## Data Availability

Health and Retirement Study, ([RAND Longitudinal]) public use dataset. Produced and distributed by the University of Michigan with funding from the National Institute on Aging (grant number NIA U01AG009740). Ann Arbor, MI, (2016).
Health and Retirement Study, ([Epigenetic Clocks]) public use dataset. Produced and distributed by the University of Michigan with funding from the National Institute on Aging (grant number NIA U01AG009740). Ann Arbor, MI, (2016).

## Acknowledgment Section

The HRS (Health and Retirement Study) is sponsored by the National Institute on Aging (grant number NIA U01AG009740) and is conducted by the University of Michigan. We acknowledge the HRS study participants and the HRS research and support staff for their contributions to this study. The content is solely the responsibility of the authors and does not necessarily represent the official views of the NIH. Jennifer Manly had full access to all the data in the study and takes responsibility for the integrity of the data and the accuracy of the data analysis.

---

**Figure 1. Socioeconomic Gradients in Memory Level and Decline Across Racial/Ethnic-Sex/Gender Subgroups**

**Figure 2. Socioeconomic Gradients in DNA-methylation Measures of Aging Across Racial/Ethnic-Sex/Gender Subgroups.**

**Figure 3. Relationship Between DNA-methylation Measures of Aging and Memory Level and Decline Across Racial/Ethnic-Sex/Gender Subgroups.**

## References

1. Campisi J, Kapahi P, Lithgow GJ, Melov S, Newman JC, Verdin E. From discoveries in ageing research to therapeutics for healthy ageing. Nature. 2019;571(7764):183–192. doi:10.1038/s41586-019-1365-2

2. Gladyshev VN. The Ground Zero of Organismal Life and Aging. Trends Mol Med. 2021;27(1):11–19. doi:10.1016/j.molmed.2020.08.012

3. Belsky DW, Caspi A, Houts R, et al. Quantification of biological aging in young adults. Proc Natl Acad Sci. 2015;112(30):E4104–E4110. doi:10.1073/pnas.1506264112

4. Elliott ML, Caspi A, Houts RM, et al. Disparities in the pace of biological aging among midlife adults of the same chronological age have implications for future frailty risk and policy. Nat Aging. 2021;1(3):295–308. doi:10.1038/s43587-021-00044-4

5. Elliott ML, Belsky DW, Knodt AR, et al. Brain-age in midlife is associated with accelerated biological aging and cognitive decline in a longitudinal birth cohort. Mol Psychiatry. Published online December 10, 2019:1-10. doi:10.1038/s41380-019-0626-7

6. Belsky DW, Moffitt TE, Cohen AA, et al. Eleven Telomere, Epigenetic Clock, and Biomarker-Composite Quantifications of Biological Aging: Do They Measure the Same Thing? Am J Epidemiol. 2018;187(6):1220–1230. doi:10.1093/aje/kwx346

7. Murman DL. The Impact of Age on Cognition. Semin Hear. 2015;36(3):111–121. doi:10.1055/s-0035-1555115

8. Steptoe A, Zaninotto P. Lower socioeconomic status and the acceleration of aging: An outcome-wide analysis. Proc Natl Acad Sci. 2020;117(26):14911–14917. doi:10.1073/pnas.1915741117

9. Duncan GJ, Magnuson K. Socioeconomic status and cognitive functioning: moving from correlation to causation. WIREs Cogn Sci. 2012;3(3):377–386. doi:https://doi.org/10.1002/wcs.1176

10. Zhang Z, Hayward MD, Yu Y-L. Life Course Pathways to Racial Disparities in Cognitive Impairment among Older Americans. J Health Soc Behav. 2016;57(2):184–199. doi:10.1177/0022146516645925

11. Greenfield EA, Moorman SM. Childhood Socioeconomic Status and Later Life Cognition: Evidence From the Wisconsin Longitudinal Study. J Aging Health. 2019;31(9):1589–1615. doi:10.1177/0898264318783489

12. Meyer OL, Mungas D, King J, et al. Neighborhood Socioeconomic Status and Cognitive Trajectories in a Diverse Longitudinal Cohort. Clin Gerontol. 2018;41(1):82–93. doi:10.1080/07317115.2017.1282911

13. Krieger N. Embodying Inequality: A Review of Concepts, Measures, and Methods for Studying Health Consequences of Discrimination: Int J Health Serv. Published online June 22, 2016. doi:10.2190/M11W-VWXE-KQM9-G97Q

14. Geronimus AT, Hicken M, Keene D, Bound J. “Weathering” and Age Patterns of Allostatic Load Scores Among Blacks and Whites in the United States. Am J Public Health. 2006;96(5):826–833. doi:10.2105/AJPH.2004.060749

15. Geronimus AT, Hicken MT, Pearson JA, Seashols SJ, Brown KL, Cruz TD. Do US Black Women Experience Stress-Related Accelerated Biological Aging? Hum Nat. 2010;21(1):19–38. doi:10.1007/s12110-010-9078-0

16. Fiorito G, Polidoro S, Dugué P-A, et al. Social adversity and epigenetic aging: a multi-cohort study on socioeconomic differences in peripheral blood DNA methylation. Sci Rep. 2017;7(1):16266. doi:10.1038/s41598-017-16391-5

17. Schmidt Charles W. Environmental Factors in Successful Aging: The Potential Impact of Air Pollution. Environ Health Perspect. 127(10):102001. doi:10.1289/EHP4579

18. Braveman P, Gottlieb L. The Social Determinants of Health: It’s Time to Consider the Causes of the Causes. Public Health Rep. 2014;129(Suppl 2):19-31.

19. Belsky DW, Caspi A, Cohen HJ, et al. Impact of early personal-history characteristics on the Pace of Aging: implications for clinical trials of therapies to slow aging and extend healthspan. Aging Cell. 2017;16(4):644–651. doi:10.1111/acel.12591

20. Belsky DW, Caspi A, Arseneault L, et al. Quantification of the pace of biological aging in humans through a blood test, the DunedinPoAm DNA methylation algorithm. Hagg S, Tyler JK, Hagg S, Justice J, Suderman M, eds. eLife. 2020;9:e54870. doi:10.7554/eLife.54870

21. Hillary RF, Stevenson AJ, McCartney DL, et al. Epigenetic measures of ageing predict the prevalence and incidence of leading causes of death and disease burden. Clin Epigenetics. 2020;12(1):115. doi:10.1186/s13148-020-00905-6

22. Fiorito G, McCrory C, Robinson O, et al. Socioeconomic position, lifestyle habits and biomarkers of epigenetic aging: a multi-cohort analysis. Aging. 2019;11(7):2045–2070. doi:10.18632/aging.101900

23. McCrory C, Fiorito G, Hernandez B, et al. GrimAge Outperforms Other Epigenetic Clocks in the Prediction of Age-Related Clinical Phenotypes and All-Cause Mortality. J Gerontol Ser A. 2020;(glaa286). doi:10.1093/gerona/glaa286

24. Hillary RF, Stevenson AJ, Cox SR, et al. An epigenetic predictor of death captures multi- modal measures of brain health. Mol Psychiatry. Published online December 3, 2019:1-11. doi:10.1038/s41380-019-0616-9

25. Sonnega A, Weir D. The Health and Retirement Study: A Public Data Resource for Research on Aging. Open Health Data. 2014;2(1):e7. doi:10.5334/ohd.am

26. Ofstedal MB, Weir DR. Recruitment and Retention of Minority Participants in the Health and Retirement Study. The Gerontologist. 2011;51(Suppl 1):S8-S20. doi:10.1093/geront/gnq100

27. Fisher GG, Ryan LH. Overview of the Health and Retirement Study and Introduction to the Special Issue. Work Aging Retire. 2018;4(1):1–9. doi:10.1093/workar/wax032

28. Crimmins E, Faul J, Kim JK, Thyagarajan B, Weir D. HRS 2016 VBS – Innovative Sub Sample Assays: Homocysteine, Clusterin, Brain-derived Neurotrophic Factor (BDNF), and mtDNA Copy Number. :7.

29. Ferrucci L, Gonzalez-Freire M, Fabbri E, et al. Measuring biological aging in humans: A quest. Aging Cell. 2020;19(2):e13080. doi:https://doi.org/10.1111/acel.13080

30. Levine ME, Lu AT, Quach A, et al. An epigenetic biomarker of aging for lifespan and healthspan. Aging. 2018;10(4):573–591. doi:10.18632/aging.101414

31. Lu AT, Quach A, Wilson JG, et al. DNA methylation GrimAge strongly predicts lifespan and healthspan. Aging. 2019;11(2):303–327. doi:10.18632/aging.101684

32. Horvath S, Raj K. DNA methylation-based biomarkers and the epigenetic clock theory of ageing. Nat Rev Genet. 2018;19(6):371–384. doi:10.1038/s41576-018-0004-3

33. Tannenbaum C, Greaves L, Graham ID. Why sex and gender matter in implementation research. BMC Med Res Methodol. 2016;16(1):145. doi:10.1186/s12874-016-0247-7

34. Hurd MD, Meijer E, Moldoff M, Rohwedder S. Improved Wealth Measures in the Health and Retirement Study: Asset Reconciliation and Cross-Wave Imputation. :79.

35. Friedline T, Masa RD, Chowa GAN. Transforming wealth: Using the inverse hyperbolic sine (IHS) and splines to predict youth’s math achievement. Soc Sci Res. 2015;49:264–287. doi:10.1016/j.ssresearch.2014.08.018

36. Morris JC, Heyman A, Mohs RC, al. et. The consortium to establish a registry for Alzheimer’s disease (CERAD). Part I. Clinical and neurospychological assessment of Alzheimer’s disease. Neurology. 1989;39:1159–1165.

37. Byrd DR, Gonzales E, Moody DLB, et al. Interactive Effects of Chronic Health Conditions and Financial Hardship on Episodic Memory among Older Blacks: Findings from the Health and Retirement Study. Res Hum Dev. 2020;17(1):41–56. doi:10.1080/15427609.2020.1746159

38. Ten Have TR, Localio AR. Empirical Bayes estimation of random effects parameters in mixed effects logistic regression models. Biometrics. 1999;55(4):1022–1029. doi:10.1111/j.0006-341x.1999.01022.x

39. Mascha EJ, Dalton JE, Kurz A, Saager L. Statistical grand rounds: understanding the mechanism: mediation analysis in randomized and nonrandomized studies. Anesth Analg. 2013;117(4):980–994. doi:10.1213/ANE.0b013e3182a44cb9

40. Muthen LK, Muthen BO. Mplus User’s Guide: Statistical Analysis with Latent Variables. 3rd ed. Muthen & Muthen; 2004.

41. Yuan Y, MacKinnon DP. Bayesian mediation analysis. Psychol Methods. 2009;14(4):301–322. doi:10.1037/a0016972

42. Muthen B. Bayesian Analysis In Mplus: A Brief Introduction. Published online 2010:92.

43. du Prel J-B, Hommel G, Röhrig B, Blettner M. Confidence interval or p-value?: part 4 of a series on evaluation of scientific publications. Dtsch Arzteblatt Int. 2009;106(19):335–339. doi:10.3238/arztebl.2009.0335

44. Chen E, Miller GE. Socioeconomic status and health: mediating and moderating factors. Annu Rev Clin Psychol. 2013;9:723–749. doi:10.1146/annurev-clinpsy-050212-185634

45. Lastrucci V, Lorini C, Caini S, Group FHLR, Bonaccorsi G. Health literacy as a mediator of the relationship between socioeconomic status and health: A cross-sectional study in a population-based sample in Florence. PLOS ONE. 2019;14(12):e0227007. doi:10.1371/journal.pone.0227007

46. Deckers K, Cadar D, van Boxtel MPJ, Verhey FRJ, Steptoe A, Köhler S. Modifiable Risk Factors Explain Socioeconomic Inequalities in Dementia Risk: Evidence from a Population-Based Prospective Cohort Study. J Alzheimers Dis. 2019;71(2):549–557. doi:10.3233/JAD-190541

47. Fritz MS, MacKinnon DP. Required Sample Size to Detect the Mediated Effect. Psychol Sci. 2007;18(3):233–239. doi:10.1111/j.1467-9280.2007.01882.x

48. Justice JN, Kritchevsky SB. Putting epigenetic biomarkers to the test for clinical trials. eLife. 2020;9:e58592. doi:10.7554/eLife.58592

49. Lawrence KG, Kresovich JK, O’Brien KM, et al. Association of Neighborhood Deprivation With Epigenetic Aging Using 4 Clock Metrics. JAMA Netw Open. 2020;3(11):e2024329–e2024329. doi:10.1001/jamanetworkopen.2020.24329

50. Dawber TR, Meadors GF, Moore FE. Epidemiological approaches to heart disease: the Framingham Study. Am J Public Health Nations Health. 1951;41(3):279–281. doi:10.2105/ajph.41.3.279

51. Mohadjer L, Bell B, Waksberg J. Accounting for Item Non-Response Bias. Published online 1994:67.

