## Supplementary Methods for "Socioeconomic Status, Biological Aging, and Memory in a Diverse National Sample of Older US Men and Women"

### Supplement

#### 1. Online only text. Detailed Description of Biological Aging Measures

**Detailed Description of Biological Aging Measures**

Epigenetic clocks are algorithms that combine information from DNA methylation (DNAm) measurements from across the genome to quantify variation in biological age.[1–3] The first-generation of clocks were developed using machine-learning to predict chronological age.[2,3] Recently, a second generation of clocks were developed to predict aging related mortality risk, incorporating information from DNAm prediction of physiological parameters. These second-generation are more predictive of morbidity and mortality.[4,5]

The PhenoAge clock was developed in a 2-step process using data from the US NHANES and the Italian InCHIANTI study.[4] In the first step, machine learning was used to develop a mortality prediction algorithm from 42 blood-chemistry analytes in data from the US NHANES III. This analysis selected 9 blood biomarkers (albumin, alkaline phosphatase, C-reactive protein, creatinine, glucose, white-blood-cell count, lymphocyte %, mean cell volume, and red-cell distribution width) and chronological age to form the final model (Blood-Chemistry PhenoAge). This model was then applied to participants in the InCHIANTI study and machine learning was used to develop a DNA methylation algorithm to predict Blood-Chemistry PhenoAge.

The GrimAge clock was developed in a 2-step process using data from the Framingham Heart Study Offspring cohort.[5] In the first step, machine-learning analysis was applied to develop DNAm algorithms for 88 blood proteins, with 12 of these algorithms meeting criteria to be carried forward to the second stage. In the second step, machine-learning analysis was used to fit DNAm-predictions of blood proteins, smoking history, age, and sex to mortality data. The final model included seven of the DNAm-predicted protein levels (adrenomedullin, beta-2-microglobulin, cystatin C, growth-differentiation factor 15, leptin, plasminogen activation inhibitor 1, tissue inhibitor metalloproteinase 1) DNAm-predicted smoking pack years, chronological age, and sex.

The DunedinPoAm pace of aging measure was developed in a 2-step process using data from the Dunedin Longitudinal Study.[6] In the first step, growth modeling was used to estimate rates of change for 18 biomarkers of organ system integrity over a 12-year follow-up period spanning ages 26-38. Rates of change were composited across biomarkers to form the Pace of Aging measure. In the second step, machine-learning analysis was applied to develop a DNAm predictor of pace of aging based on DNA collected at the end of the follow-up interval.

**References**


1. Shireby GL, Davies JP, Francis PT, et al. Recalibrating the epigenetic clock: implications for assessing biological age in the human cortex. *Brain*. 2020;143(12):3763-3775. doi:10.1093/brain/awaa334

2. Hannum G, Guinney J, Zhao L, et al. Genome-wide methylation profiles reveal quantitative views of human aging rates. *Mol Cell*. 2013;49(2):359-367. doi:10.1016/j.molcel.2012.10.016

3. Horvath S. DNA methylation age of human tissues and cell types. *Genome Biology*. 2013;14(10):3156. doi:10.1186/gb-2013-14-10-r115

4. Levine ME, Lu AT, Quach A, et al. An epigenetic biomarker of aging for lifespan and healthspan. *Aging (Albany NY)*. 2018;10(4):573-591. doi:10.18632/aging.101414

5. Lu AT, Quach A, Wilson JG, et al. DNA methylation GrimAge strongly predicts lifespan and healthspan. *Aging (Albany NY)*. 2019;11(2):303-327. doi:10.18632/aging.101684

6. Belsky DW, Caspi A, Arseneault L, et al. Quantification of the pace of biological aging in humans through a blood test, the DunedinPoAm DNA methylation algorithm. Hagg S, Tyler JK, Hagg S, Justice J, Suderman M, eds. *eLife*. 2020;9:e54870. doi:10.7554/eLife.54870
